# Respiratory morbidity in preschool and school-age children born very preterm and its association with parents’ health-related quality of life and family functioning

**DOI:** 10.1101/2022.08.10.22278622

**Authors:** Gabriela P. Peralta, Raffaela Piatti, Sarah R. Haile, Mark Adams, Dirk Bassler, Alexander Moeller, Giancarlo Natalucci, Susi Kriemler

## Abstract

**Importance:** It is not known whether respiratory morbidity associated with very premature birth and bronchopulmonary dysplasia (BPD) influence parents’ health and family functioning beyond infancy.

**Objective:** To describe the prevalence and severity of respiratory symptoms in children born very preterm and to assess their association with parents’ health-related quality of life (HRQoL) and family functioning.

**Design, setting and participants:** In this cross-sectional study, we recruited children born less than 32 weeks’ gestation between January 2006 and December 2019, in the greater Zurich area, Switzerland. Between May and December 2021, parents were invited to complete an online survey for their preterm child and for a control term born (≥37 weeks’ gestation) sibling aged 1 to 18 years. Children with severe chronic conditions or with a recent COVID-19 infection were excluded.

**Exposure:** Respiratory symptoms, categorised as ‘none’, ‘mild’ and ‘moderate-severe’.

**Main outcomes and measures:** The Total Score, the Parent HRQoL Score, and the Family Functioning Score of the Pediatrics Quality of Life Family Impact Module (PedsQL FIM) questionnaire. Associations between respiratory symptoms and these scores were assessed using multivariable linear regression, adjusted for potential confounders.

**Results:** Of 1,697 eligible very preterm children, the survey was completed for 616 of them (99 with BPD) and 180 controls. Girls made up 45% (46% in controls) of the sample and 63% (60% in controls) of participants were aged 6 to 18 years (school-age). Overall, very preterm children reported a higher risk of respiratory symptoms than controls, especially the preschool group and those with moderate-to-severe BPD. Parents of children with ‘mild’ and ‘moderate-severe’ respiratory symptoms had on average -3.9 [95%CI: -6.6 to -1.1] and -8.2 [-11.2 to -5.2] lower Total Score respectively than parents of children with no symptoms. The same pattern was observed for the other summary scores, and after stratifying by age categories.

**Conclusions and relevance:** Our study suggests that respiratory morbidity in very preterm children has a negative impact on parents’ HRQoL and family functioning, even beyond the first years of life. This finding highlights the need for appropriate monitoring and support for families of survivors of very premature birth.

**KEY POINTS:** *Question:* Is respiratory morbidity in survivors of very premature birth associated with parents’ health-related quality of life (HRQoL) and family functioning beyond the first years of life?

*Findings:* In this cross-sectional study including 616 children born very preterm and 185 term born controls, both aged 1 to 18 years, we found that very preterm children remain at increased risk of respiratory morbidity through childhood, and that this is associated with decreased parents’ HRQoL and family functioning.

*Meaning:* Families of survivors of very premature birth need appropriate monitoring and support to cope with the burden of respiratory morbidity.

## BACKGROUND

Over the last decades, the prevalence of very preterm birth (<32 completed weeks’ gestation) has risen substantially.^1^ Advances in neonatal care have improved survival rates,^2,3^ but infants who survive are likely to face serious neonatal morbidity. The lungs of very preterm infants are not fully developed at birth and therefore are susceptible to suboptimal further development and to lung injury due to neonatal factors, such as mechanical ventilation or oxygen therapy.^4,5^ Indeed, despite advances in respiratory management, the prevalence of bronchopulmonary dysplasia (BPD), a neonatal chronic lung disease, has not decreased.^6,7^ Previous research has shown that the burden of respiratory morbidity associated with prematurity and BPD are high and last far beyond the neonatal period.^8^ Premature survivors, with or without BPD, require re-hospitalization and inhalation therapy more often than full-term children during the first year of life.^9^ Also, compared to term born children, very preterm children have a higher risk of wheezing disorders^10^ and show altered lung function trajectories in childhood, particularly those with BPD.^11^

The persistence of respiratory symptoms throughout childhood in survivors of very premature birth may place an important burden on their parents and families, accentuating the burden that they already bear due to prematurity per se and other complications associated with it. Compared to parents of term born children, parents of premature children have an increased risk of psychological distress and are likely to report higher levels of stress and poorer family functioning during infancy and early childhood.^12–14^ In addition, previous studies have shown that BPD and respiratory symptoms contribute to lower health-related quality of life (HRQoL) in parents of very preterm children during the first years of life.^15–17^ However, despite this evidence, no previous study has assessed whether respiratory morbidity in survivors of very premature birth continues to affect parents’ health and family functioning beyond infancy.

In this study, we aimed to describe the prevalence and severity of respiratory symptoms in preschool and school-age children born very preterm in comparison to a sample of children born at term, and to assess the associations of respiratory morbidity in very preterm children with parents’ HRQoL and family functioning.

## METHODS

Complete details are provided in the Supplement.

### Study design and participants

In this cross-sectional study we recruited children born less than 32 weeks’ gestation between January 2006 and December 2019, in the greater Zurich area, Switzerland. They were all included in the Swiss Neonatal Network & Follow-Up Group (SwissNeoNet), a nationwide registry of very preterm children.^18^ Of 1,697 eligible children, valid postal addresses were obtained for 1,401 of them. Invitation letters were sent out in six rounds between May and December 2021 and included a detailed description of the study procedures in plain language. Parents were invited to complete an online survey for their preterm child as well as for a term born (≥37 weeks’ gestation) sibling aged 1 to 18 years, referred as controls hereafter. Families who did not complete the survey within two weeks received a reminder call or a second invitation letter, if the phone number was not available. Parents were given the option to complete a paper version and with translation into English, French and Italian. We finalized data collection in May 2022.

After exclusion of children with severe chronic conditions or with a recent COVID-19 infection, 616 very preterm children (44% of the invited) and 180 controls were included in the study (see details in the Supplementary Methods). The analysis on the association between respiratory morbidity and parents’ HRQoL was restricted to very preterm children and for families with twins or triplets one child was randomly selected (n=533, Figure S1 in the Supplement).

The study was approved by the Ethics Committee of the Canton of Zurich, Switzerland (2020-02396). Filling out the online survey was considered as providing consent. This study followed the Strengthening the Reporting of Observational Studies in Epidemiology (STROBE) reporting guidelines for cross-sectional studies.

### Respiratory symptoms

Parents completed a questionnaire about their child’s (children’s) respiratory health. The questionnaire included validated questions from the Swiss Paediatric Airway Cohort (SPAC) study.^19^ Respiratory symptoms were categorized into ‘mild’ and ‘moderate-severe’. Children were classified as having ‘mild’ symptoms if they reported at least one of the following: breathing difficulties during exertion, cough without cold, nocturnal cough in the past 12 months or wheezing in the past 12 months. Children were classified as having ‘moderate-severe’ symptoms if they reported at least one of the following in the past 12 months: emergency visits to the pediatrician or family doctor, a visit to a hospital’s emergency ward, a hospitalization or inhalation therapy for respiratory problems. We combined ‘mild’ and ‘moderate-severe’ symptoms into a single variable with three categories: none, mild (only mild symptoms reported) and moderate-severe (both mild and moderate-severe symptoms, or only moderate-severe symptoms reported). This combined variable was used as the main exposure variable to assess the association between respiratory morbidity and parents’ HRQoL and family functioning.

The questions used to assess each respiratory symptom are presented in Tables S1 in the Supplement.

### Parents’ health-related quality of life (HRQoL) and family functioning

The Pediatrics Quality of Life Family Impact Module 2.0 (PedsQL FIM) was used to assess parents’ HRQoL and family functioning. The PedsQL FIM is a standardized self-report questionnaire that measures the impact of chronic pediatric health conditions on parental quality of life and family functioning with reference to the last four weeks.^20,21^ It consists of 36 Likert-scaled items standing for several functional dimensions and generates three scores (ranging from 0 to 100): the Total Score, the Parent HRQoL Summary Score, and the Family Functioning Summary Score.^21^ Higher scores denote better parental HRQoL and family functioning.

### Covariates

Neonatal characteristics for very preterm children were retrieved from the SwissNeoNet registry. Gestational age was categorized as 22 to 28 weeks (extremely preterm) or 28 to 31 weeks (very preterm). BPD status was defined as oxygen use for 28 days and its severity was defined based on oxygen dependency at 36 weeks post-menstrual age.^22^ Premature children were classified as no-to-mild BPD and moderate-to-severe BPD.^22^ Birth weight z-scores and socioeconomic status of the family were defined as previously published.^23^

We collected information on children’s sex, age, weight, and height in the online survey. We used height and weight to derive age- and sex-specific body mass index (BMI) z-scores.^24^ We also collected information on maternal smoking during pregnancy, parents’ nationality, educational level, smoking status, history of atopy, number of siblings, presence of chronic conditions and who filled in the questionnaire. We classified children’s age as 1 to 5 years (preschool age) and 6 to 18 years (school age).

### Statistical Analysis

Data were presented as frequencies with percentages or as medians with interquartile ranges. Descriptive analyses for respiratory symptoms were stratified by age and BPD categories. Between group comparisons were assessed using unadjusted logistic regression.

To study the associations between the respiratory symptoms in very preterm children and the PedsQL FIM scores, we used multivariable linear regression. Models were adjusted for child’s sex, age category, gestational age categories, chronic conditions, number of siblings, parent’s nationality, educational level, smoking status, history of atopy and for who filled in the survey. Models were performed for the overall sample and stratified by age category. To avoid biases due to missing data, we implemented multiple imputation by chained equations for missing values of exposure, outcomes and covariates, generating 25 complete datasets. The percentage of missing data for each variable is presented in Table S2 in the Supplement.

We performed several sensitivity analyses to assess the robustness of our findings. We repeated the models after excluding parents with PedsQL FIM scores below the first percentile (to assess the influence of potential outliers) and using the observed data (i.e., without imputation). Finally, we repeated the models in the observed data by using 100 different random samples for twins and triplets. All analyses were conducted using the statistical software R (version 4.2.0).

## RESULTS

### Study sample

The characteristics of the study sample are presented in Table 1. Control and very preterm groups were comparable in terms of sex, age, BMI z-score and sociodemographic characteristics. Very preterm children had a median (P_25_; P_75_) gestational age of 29 weeks (27; 30) and 99 (16%) of them were classified as having moderate-to-severe BPD. Parents of participants were mostly Swiss and most of them had a high education level. Compared to very premature children that did not participate in the study, participants had a lower gestational age, lower birth weight, were more likely to have moderate-to-severe BPD and were from a higher socioeconomic status (Table S3 in the Supplement).

**Table 1.**
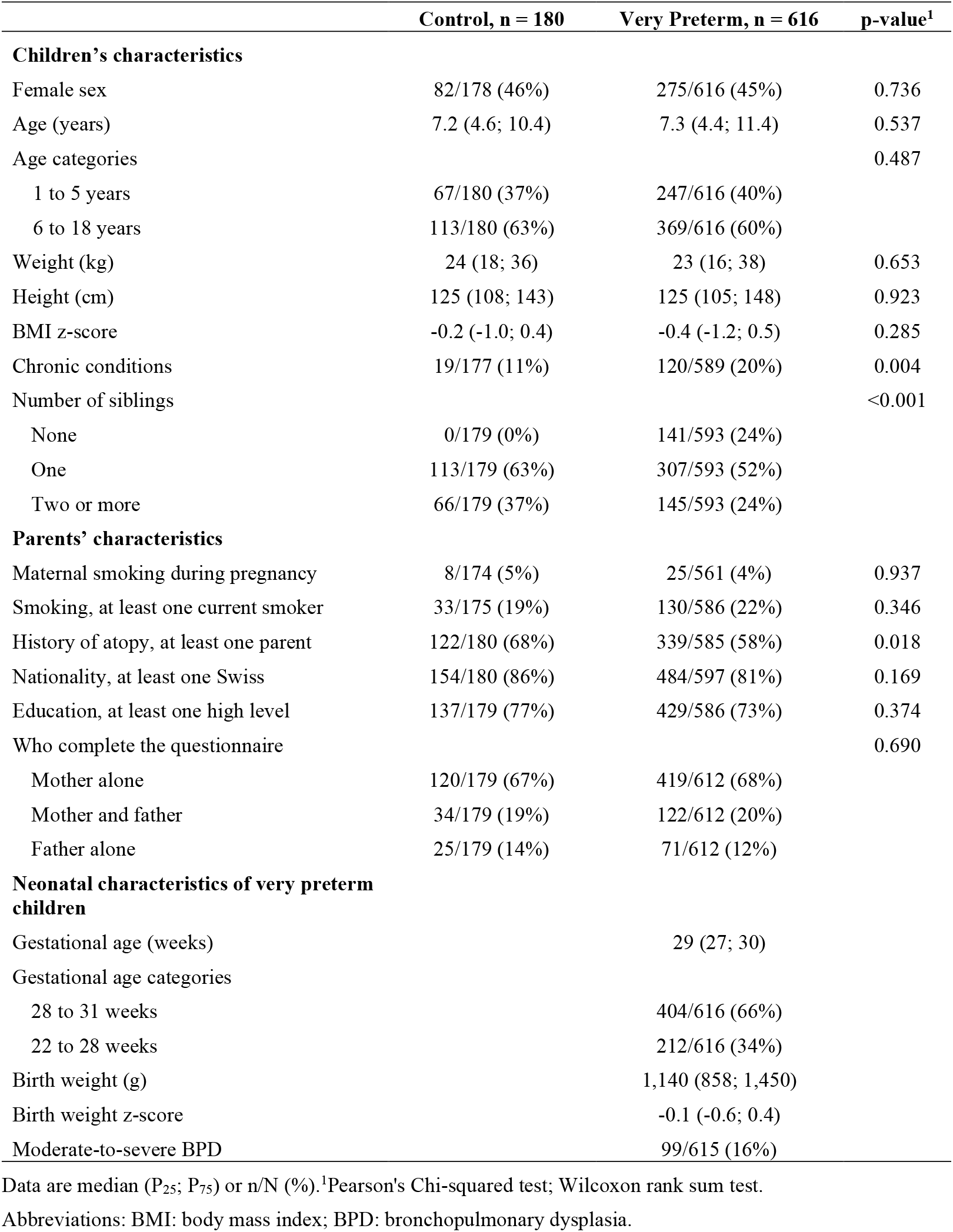
Characteristics of the study sample.

|  | Control, n = 180 | Very Preterm, n = 616 | p-value<sup>1</sup> |
| --- | --- | --- | --- |
| **Children's characteristics** |  |  |  |
| Female sex | 82/178 (46%) | 275/616 (45%) | 0.736 |
| Age (years) | 7.2 (4.6; 10.4) | 7.3 (4.4; 11.4) | 0.537 |
| Age categories |  |  | 0.487 |
| 1 to 5 years | 67/180 (37%) | 247/616 (40%) |  |
| 6 to 18 years | 113/180 (63%) | 369/616 (60%) |  |
| Weight (kg) | 24 (18; 36) | 23 (16; 38) | 0.653 |
| Height (cm) | 125 (108; 143) | 125 (105; 148) | 0.923 |
| BMI z-score | -0.2 (-1.0; 0.4) | -0.4 (-1.2; 0.5) | 0.285 |
| Chronic conditions | 19/177 (11%) | 120/589 (20%) | 0.004 |
| Number of siblings |  |  | <0.001 |
| None | 0/179 (0%) | 141/593 (24%) |  |
| One | 113/179 (63%) | 307/593 (52%) |  |
| Two or more | 66/179 (37%) | 145/593 (24%) |  |
| **Parents' characteristics** |  |  |  |
| Maternal smoking during pregnancy | 8/174 (5%) | 25/561 (4%) | 0.937 |
| Smoking, at least one current smoker | 33/175 (19%) | 130/586 (22%) | 0.346 |
| History of atopy, at least one parent | 122/180 (68%) | 339/585 (58%) | 0.018 |
| Nationality, at least one Swiss | 154/180 (86%) | 484/597 (81%) | 0.169 |
| Education, at least one high level | 137/179 (77%) | 429/586 (73%) | 0.374 |
| Who complete the questionnaire |  |  | 0.690 |
| Mother alone | 120/179 (67%) | 419/612 (68%) |  |
| Mother and father | 34/179 (19%) | 122/612 (20%) |  |
| Father alone | 25/179 (14%) | 71/612 (12%) |  |
| **Neonatal characteristics of very preterm children** |  |  |  |
| Gestational age (weeks) |  | 29 (27; 30) |  |
| Gestational age categories |  |  |  |
| 28 to 31 weeks |  | 404/616 (66%) |  |
| 22 to 28 weeks |  | 212/616 (34%) |  |
| Birth weight (g) |  | 1,140 (858; 1,450) |  |
| Birth weight z-score |  | -0.1 (-0.6; 0.4) |  |
| Moderate-to-severe BPD |  | 99/615 (16%) |  |

Data are median (P<sub>25</sub>; P<sub>75</sub>) or n/N (%).<sup>1</sup>Pearson's Chi-squared test; Wilcoxon rank sum test.
Abbreviations: BMI: body mass index; BPD: bronchopulmonary dysplasia.

### Respiratory symptoms

Table 2 and Table 3 show the prevalence of respiratory symptoms in preschool and school-age children, respectively. In the preschool-age group, very preterm children with no-to-mild BPD and with moderate-to-severe BPD were more likely to present ‘mild’ symptoms (unadjusted odds ratio [95% CI]: 3.1 [1.6 to 6.1], *P*<0.001 and 4.1 [1.8 to 9.6], *P*<0.001, respectively) than controls. Differences were especially relevant for breathing difficulties during exertion and nocturnal cough in the past 12 months. Compared to controls, very preterm children with moderate-to-severe BPD also appeared to be more likely to present ‘moderate-severe’ symptoms (2.4 [1.0 to 6.1], *P*=0.053), while this was not the case for very preterm children with no-to-mild BPD. In the school-age group, compared to controls, very preterm children with no-to-mild BPD were more likely to report breathing difficulties during exertion but there was no evidence for differences in the prevalence of other respiratory symptoms. In contrast, very preterm children with moderate-to-severe BPD appeared to be more likely to have ‘mild’ (2.0 [0.9 to 4.2], *P*=0.090) and ‘moderate-severe’ (2.4 [0.6 to 6.5], *P=*0.093) symptoms than controls, although evidence was weak. This group was also more likely to report breathing difficulties during exertion and inhalation therapy in the past 12 months than controls. We found no evidence for differences in the prevalence of respiratory symptoms according to BPD categories in very preterm born children, in either age group.

**Table 2.** Prevalence of mild and moderate-severe respiratory symptoms and between group comparisons in preschool-age children.

| Symptoms | C,<br>n = 67 | VP no BPD,<br>n = 191 | VP BPD,<br>n = 55 | VP no BPD vs. C | VP BPD vs. C | VP BPD vs. VP no<br>BPD |
| --- | --- | --- | --- | --- | --- | --- |
| **Mild symptoms** | 14/65 (22%) | 81/177 (46%) | 25/47 (53%) | 3.1 [1.6 to 6.1] | 4.1 [1.8 to 9.6] | 1.3 [0.7 to 2.6] |
| Breathing difficulties during exertion | 1/64 (2%) | 16/176 (9%) | 7/49 (14%) | 6.3 [1.2 to 114.9] | 10.5 [1.8 to 200.0] | 1.7 [0.6 to 4.2] |
| Cough without cold | 6/67 (9%) | 31/191 (16%) | 14/52 (27%) | 2.0 [0.8 to 5.4] | 3.7 [1.4 to 11.4] | 1.9 [0.9 to 3.9] |
| Nocturnal cough past 12 months | 3/66 (5%) | 38/183 (21%) | 11/46 (24%) | 5.5 [1.9 to 23.4] | 6.6 [1.9 to 30.6] | 1.2 [0.5 to 2.5] |
| Wheezing past 12 months | 7/65 (11%) | 35/185 (19%) | 11/51 (22%) | 1.9 [0.9 to 5.0] | 2.3 [0.8 to 6.7] | 1.2 [0.5 to 2.5] |
| **Moderate-severe symptoms** | 10/63 (16%) | 43/164 (26%) | 16/51 (31%) | 1.9 [0.9 to 4.2] | 2.4 [1.0 to 6.1] | 1.3 [0.6 to 2.5] |
| Emergency visits or hospitalization past 12 months | 6/62 (10%) | 31/165 (19%) | 13/51 (25%) | 2.2 [0.9 to 6.0] | 3.2 [1.2 to 9.8] | 1.5 [0.7 to 3.1] |
| Inhalation therapy past 12 months | 8/67 (12%) | 29/179 (16%) | 12/53 (23%) | 1.4 [0.6 to 3.5] | 2.2 [0.8 to 6.0] | 1.5 [0.7 to 3.2] |

Data are presented as n/N (%). Between group comparisons are presented as unadjusted OR (95% CI). p-values for between group comparisons are presented in Table S5
Abbreviations: BPD, bronchopulmonary dysplasia; C, controls; CI, confidence interval; OR, odds ratio; VP no BPD, very preterm with no-to-mild BPD; VP BPD, very preterm with moderate-to-severe BPD.

**Table 3.** Prevalence of mild and moderate-severe respiratory symptoms and between group comparisons in school-age children.

| Symptoms | C,<br>n = 113 | VP no BPD,<br>n = 325 | VP BPD,<br>n = 44 | VP no BPD vs. C | VP BPD vs. C | VP BPD vs. VP no<br>BPD |
| --- | --- | --- | --- | --- | --- | --- |
| **Mild symptoms** | 30/107 (28%) | 100/285 (35%) | 16/37 (43%) | 1.4 [0.9 to 2.3] | 2.0 [0.9 to 4.2] | 1.4 [0.7 to 2.8] |
| Breathing difficulties during exertion | 6/106 (6%) | 42/297 (14%) | 7/41 (17%) | 2.7 [1.2 to 7.4] | 3.4 [1.1 to 11.3] | 1.2 [0.5 to 2.9] |
| Cough without cold | 21/112 (19%) | 56/323 (17%) | 7/44 (16%) | 0.9 [0.5 to 1.6] | 0.8 [0.3 to 2.0] | 0.9 [0.4 to 2.0] |
| Nocturnal cough past 12 months | 11/109 (10%) | 35/288 (12%) | 5/38 (13%) | 1.2 [0.6 to 2.6] | 1.3 [0.4 to 4.0] | 1.1 [0.4 to 2.8] |
| Wheezing past 12 months | 6/109 (6%) | 32/310 (10%) | 3/43 (7%) | 2.0 [0.9 to 5.4] | 1.3 [0.3 to 5.1] | 0.7 [0.2 to 1.9] |
| **Moderate-severe symptoms** | 10/108 (9%) | 40/301 (13%) | 8/41 (20%) | 1.5 [0.7 to 3.3] | 2.4 [0.8 to 6.5] | 1.6 [0.6 to 3.5] |
| Emergency visits or hospitalization past 12 months | 4/108 (4%) | 17/303 (6%) | 4/42 (10%) | 1.5 [0.6 to 5.5] | 2.7 [0.6 to 12.1] | 1.8 [0.5 to 5.1] |
| Inhalation therapy past 12 months | 7/113 (6%) | 35/315 (11%) | 8/41 (20%) | 1.9 [0.9 to 4.8] | 3.7 [1.2 to 11.2] | 1.9 [0.8 to 4.4] |

Data are presented as n/N (%). Between group comparisons are presented as unadjusted OR (95% CI). p-values for between group comparisons are presented in Table S6
Abbreviations: BPD, bronchopulmonary dysplasia; C, controls; CI, confidence interval; OR, odds ratio; VP no BPD, very preterm with no-to-mild BPD; VP BPD, very preterm with moderate-to-severe BPD.

In the preschool group, more than 50% of very preterm children reported respiratory symptoms, compared to 28% of controls (Figure 2 and Table S7 in the Supplement). Also, they were more likely to report ‘moderate-severe’ symptoms than controls. We observed a similar pattern for school-age children, but in this case, there was no evidence for differences in the combination of respiratory symptoms between very preterm children and controls (Figure 2 and Table S7 in the Supplement).

**Figure 1.**
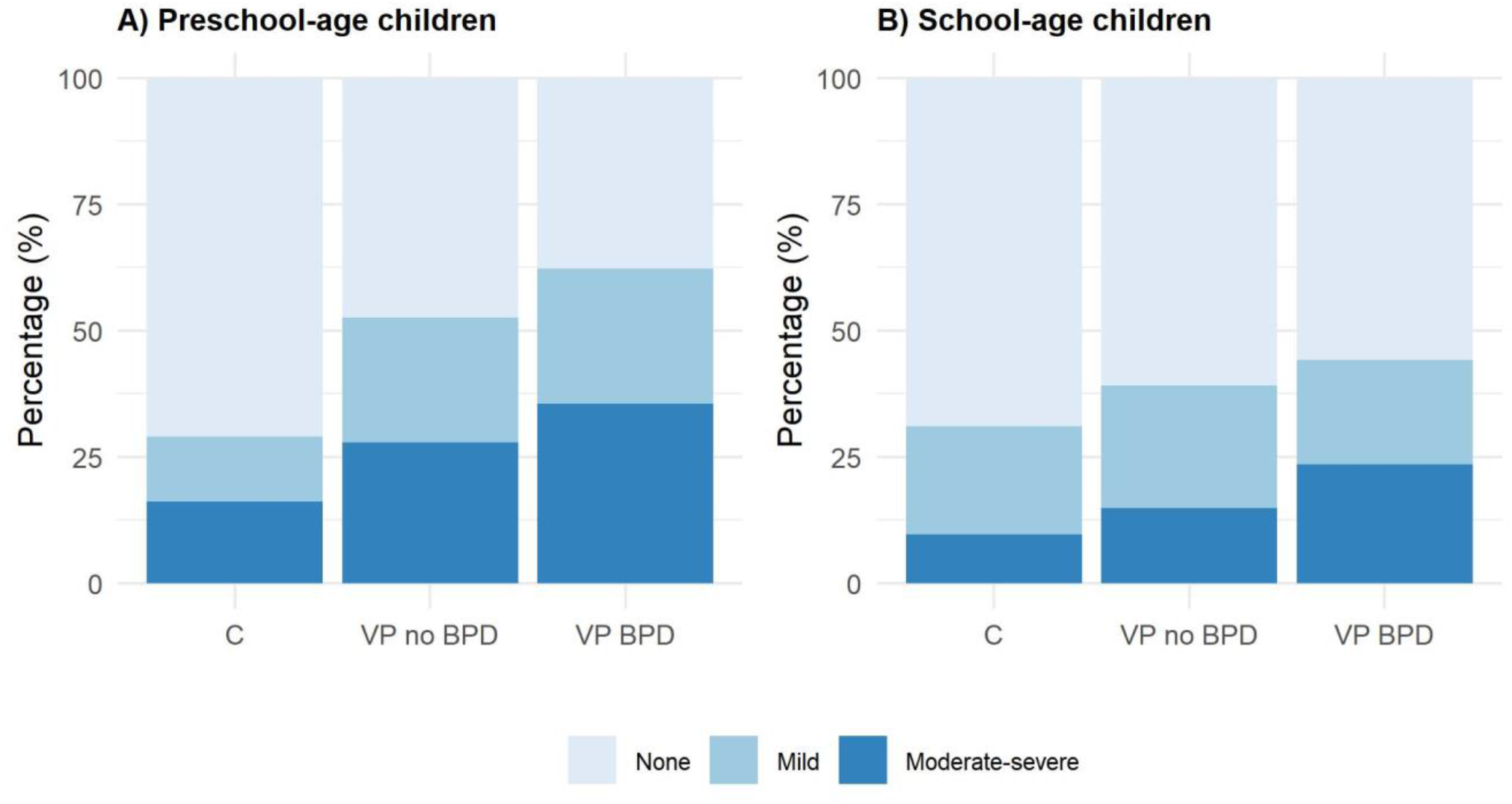
Combined variable for respiratory symptoms, stratified by age and BPD categories. Group comparisons were tested using Fisher’s exact test: preschool-age children, p-value=0.007; school-age children, p-value=0.284 Abbreviations: BPD, bronchopulmonary dysplasia; C, controls; VP no BPD, very preterm with no-to-mild BPD; VP BPD, very preterm with moderate-to-severe BPD

**Figure 2.**
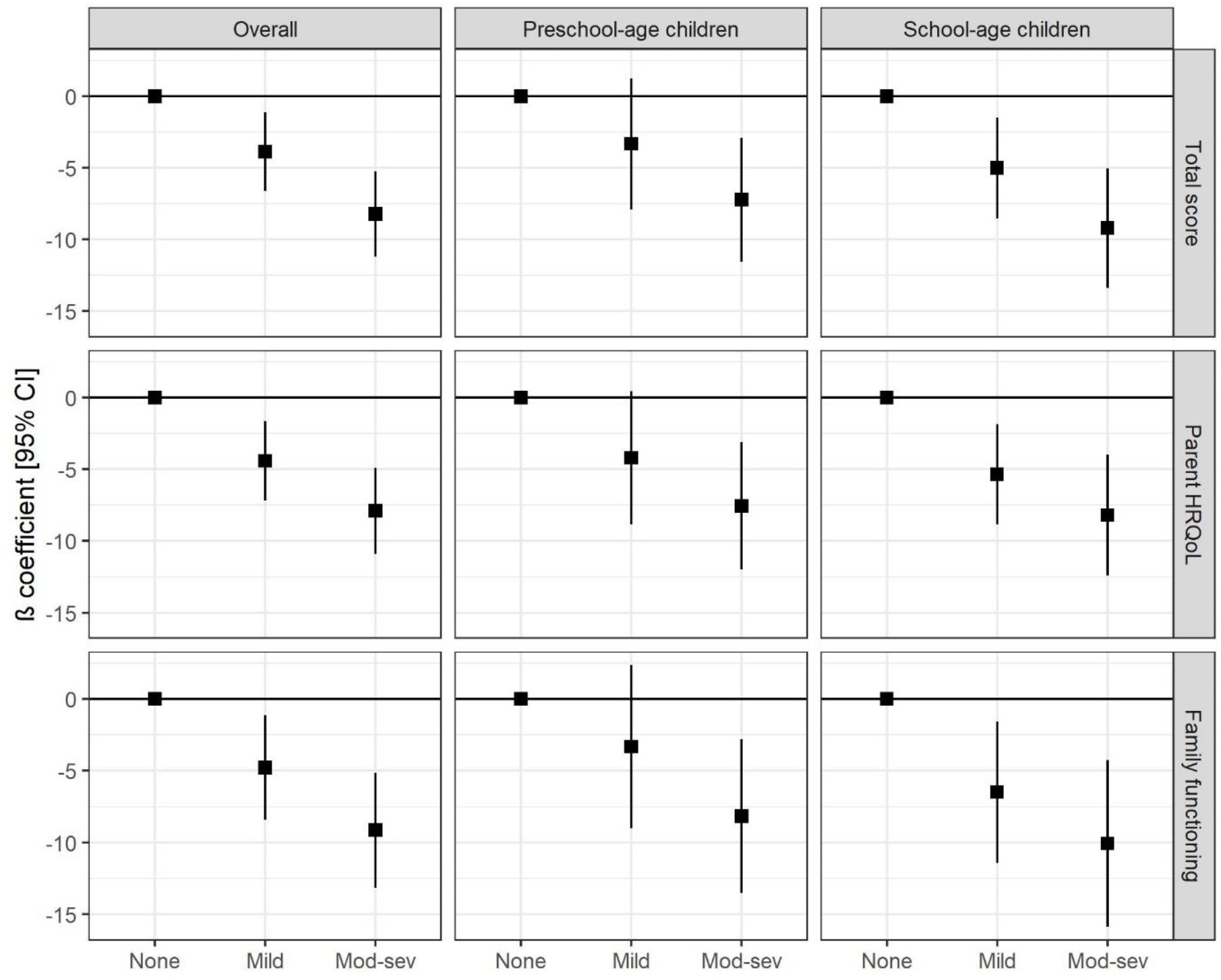
Adjusted associations between respiratory symptoms in very preterm children and parents’ HRQoL and family functioning. Abbreviations: CI, confidence interval; HRQoL, health-related quality of life; Mod-sev: moderate-severe. Models were adjusted for child’s sex, gestational age category, presence of chronic diseases, number of siblings, parents’ nationality, educational level, smoking status, family history of atopy and for who filled in the survey.

### Association of respiratory morbidity in very preterm children with parents’ HRQoL and family functioning

Table S7 in the Supplement shows descriptive statistics for the PedsQL FIM scores. Overall median scores (P_25_; P_75_) were 98.6 (88.9; 100) for the Total Score, 98.8 (88.8; 100) for the Parent HRQoL Summary Score and 100 (84.4; 100) for the Family Functioning Summary Score. Parents of preschool-age children reported lower scores than parents of school-age children.

After adjusting for relevant confounders, respiratory morbidity in very preterm children was associated with decreased parents’ HRQoL and family functioning (Figure 2 and Table S8 in the Supplement). Parents of children in the ‘mild’ and ‘moderate-severe’ categories had on average -3.9 [95%CI: -6.6 to -1.1; *P*=0.006] and -8.2 [-11.2 to -5.2; *P*<0.001] lower Total Score respectively than parents of children with no respiratory symptoms. The same pattern was observed for the other two scores. In models stratified by age category, the direction of the observed associations remained stable. However, in the group of preschool-age children, the evidence for an association between the ‘mild’ category and the Parent HRQoL Summary Score was weak and there was little evidence for an association with the Total Score and the Family Functioning Summary Score. Results were similar in all sensitivity analyses (Tables S9-S11 in the Supplement).

## DISCUSSION

In this cross-sectional study we found that very preterm children remain at increased risk of respiratory morbidity through childhood, especially those with moderate-to-severe BPD. We also found that respiratory morbidity in very preterm children is associated with decreased parental HRQoL and family functioning, both in preschool and school-age children.

Our results showing that very premature birth and BPD are associated with increased risk of respiratory symptoms through childhood are consistent with existing literature.^8,25–30^ We found that differences in respiratory morbidity were greater for preschool age children than for older children. This finding is in line with a previous study that followed children born extremely preterm during the first decade of life and reported a substantial decrease in the prevalence of respiratory symptoms and hospital admissions after five years of age.^27^ The reduction of respiratory morbidity from the start of middle childhood onwards could be attributed to airway repair and “catch-up” of alveolar growth.^31,32^ However, although respiratory symptoms appeared to decrease with age, we found evidence that school-age children born very preterm were also at increased risk for some respiratory symptoms. Specifically, compared to term born children, those born very preterm were almost three times more likely to report breathing difficulties during exertion and those with moderate-to-severe BPD were also more likely to have received inhalation therapy in the past 12 months. These findings are also in line with previous research^26,27,29^ and support the hypothesis that very premature birth and BPD have long-term consequences on respiratory health.

We found no evidence for differences in respiratory symptoms between very preterm children with no-to-mild BPD and with moderate-to-severe BPD. Although similar results have been reported previously,^30,33^ this finding contrasts with other studies that showed differences in respiratory symptoms and/or lung function according to BPD severity.^28,29,34^ Conflicting results may be due to different sample size, age range and classification of BPD. In addition, it is possible that the current definition of BPD^22^ does not allow to capture risk factors for long-term respiratory morbidity in survivors of very premature birth. In fact, a recent study suggested that gestational age and fetal growth restriction are better predictors of future lung function deficits than a history of BPD in prematurely born children.^35^

Importantly, our study suggests that respiratory morbidity in very preterm children is associated with decreased parents’ HRQoL and family functioning beyond early infancy. Parents of children with ‘moderate-severe’ respiratory symptoms were the ones who reported lower scores, both in preschool and school-age children. Previous studies have shown that premature birth has a long-term negative impact on parental and family outcomes^36,37^ and that parents of children born very preterm remain at high risk of depression and anxiety several years after birth, even after accounting for child neurodevelopmental disability.^37^ Our findings suggest that respiratory morbidity may contribute to this long-term association. Families of children with more severe respiratory symptoms are likely to face greater healthcare costs and disruption of their daily life.^38,39^ Also, the presence of respiratory health problems may entail special care needs that could adversely affect parental job stability and time for personal care and leisure activities,^40,41^ which, in turn, may have a negative impact on their well-being and family dynamics. We found that even the presence of ‘mild’ symptoms alone related to decreased parents’ HRQoL and family functioning in school-age children, while this association was less strong for preschool-age children. Respiratory morbidity in school-age children could result in more missed school days and, in consequence, in a greater disruption of work-family balance.^42^

This study has important implications for public health. Our findings, together with previous research, show that respiratory morbidity associated with very premature birth is likely to persist far beyond the neonatal period. This highlights the indisputable need of long-term monitoring strategies for survivors of very premature birth, especially for those with a history of moderate-to-severe BPD, to ensure that they receive appropriate treatment. Several studies have shown that uncontrolled respiratory symptoms in childhood can have long lasting effects on lung function growth.^43–45^ The negative association between respiratory morbidity and parents’ HRQoL also requires special attention. Previous research has reported that parental quality of life and mental health play a crucial role on the control and course of children’s respiratory illness.^46^ Therefore, this perspective is essential both for a more extensive understanding of respiratory health in children and potential interventions.

### Strengths and limitations

Strengths of this study include the detailed assessment of respiratory symptoms and the broad age range of participants, which allowed us to assess the prevalence of respiratory symptoms and its association with parents’ HRQoL and family functioning separately for pre-school and school-age children. The selection of controls among the term born siblings of children born very preterm is another strength of the study, since siblings share the same environment and the atopic family constellation that could potentially affect respiratory symptoms.

The cross-sectional design and the small sample size for some of the groups, which may have diminished the power to identify between-group differences for respiratory symptoms, are limitations of the study. Also, the limited response rate and resulting potential selection bias may affect our results: very preterm children participating in the study had a lower gestational age, lower birth weight, were more likely to have moderate-to-severe BPD and were from a higher socioeconomic status than eligible children that did not participate in the study. In addition, the assessment of respiratory symptoms was based on parental report and therefore it may be affected by potential recall bias. Also, parents of older children may be less aware of their children’s respiratory symptoms as older children are more independent. Finally, although the analysis assessing the association of respiratory symptoms with parents’ HRQoL and family functioning was adjusted for a wide range of potential confounders (including children’s chronic conditions and family history of atopy), we cannot rule out potential residual confounding.

### Conclusion

Our study, together with existent literature, provide evidence that survivors of very premature birth remain at increased risk of respiratory morbidity through childhood, especially those with a history of moderate-to-severe BPD. Importantly, our study suggests that respiratory morbidity in very preterm children has a negative impact on parents’ HRQoL and family functioning, even beyond the first years of life. This finding highlights the need for appropriate monitoring and support for families of survivors of very premature birth.

## Data Availability

Reasonable requests to share data will be subject to institutional agreements and ethics approvals. Data requests should be sent by email to the corresponding author.

## Author Contributions

Dr Peralta and Dr Kriemler had full access to all the data in the study and take responsibility for the integrity of the data and the accuracy of the data analysis. Dr Peralta and Ms Piatti share first authorship.

*Concept and design:* All authors.

*Acquisition, analysis, or interpretation of data*: All authors.

*Drafting of the manuscript:* Peralta, Piatti.

*Critical revision of the manuscript for important intellectual content:* All authors.

*Statistical Analysis:* Peralta, Haile.

*Obtained funding:* Peralta, Kriemler

*Administrative, technical, or material support:* Peralta, Piatti, Adams, Kriemler

*Supervision*: Kriemler.

## Additional Contributions

We acknowledge further contributors: Ajay Bharadwaj, Danielle Isler, and Manuel Stähli for their support in contacting families, organizational work, and data management. We are extremely grateful to all parents and children for their willingness to participate in the study. We thank the SPAC team for their collaboration regarding their questionnaire.

## REFERENCES

1. Blencowe H, Cousens S, Oestergaard MZ, et al. National, regional, and worldwide estimates of preterm birth rates in the year 2010 with time trends since 1990 for selected countries: A systematic analysis and implications. Lancet. 2012;379(9832):2162–2172. doi:10.1016/S0140-6736(12)60820-4

2. Horbar JD, Carpenter JH, Badger GJ, et al. Mortality and Neonatal Morbidity Among Infants 501 to 1500 Grams From 2000 to 2009. Pediatrics. 2012;129(6):1019–1026. doi:10.1542/peds.2011-3028

3. Stoll BJ, Hansen NI, Bell EF, et al. Trends in Care Practices, Morbidity, and Mortality of Extremely Preterm Neonates, 1993-2012. JAMA. 2015;314(10):1039. doi:10.1001/jama.2015.10244

4. Gibbons JTD, Wilson AC, Simpson SJ. Predicting Lung Health Trajectories for Survivors of Preterm Birth. Front Pediatr. 2020;8:318. doi:10.3389/fped.2020.00318

5. Smith LJ, McKay KO, van Asperen PP, Selvadurai H, Fitzgerald DA. Normal development of the lung and premature birth. Paediatr Respir Rev. 2010;11(3):135–142. doi:10.1016/j.prrv.2009.12.006

6. Costeloe KL, Hennessy EM, Haider S, Stacey F, Marlow N, Draper ES. Short term outcomes after extreme preterm birth in England: Comparison of two birth cohorts in 1995 and 2006 (the EPICure studies). BMJ. 2012;345(7886):1–14. doi:10.1136/bmj.e7976

7. Thébaud B, Goss KN, Laughon M, et al. Bronchopulmonary dysplasia. Nat Rev Dis Prim. 2019;5(1):78. doi:10.1038/s41572-019-0127-7

8. Islam JY, Keller RL, Aschner JL, Hartert TV, Moore PE. Understanding the short- and long-term respiratory outcomes of prematurity and bronchopulmonary dysplasia. Am J Respir Crit Care Med. 2015;192(2):134–156. doi:10.1164/rccm.201412-2142PP

9. Pramana I, Latzin P, Schlapbach L, et al. Respiratory symptoms in preterm infants: burden of disease in the first year of life. Eur J Med Res. 2011;16(5):223. doi:10.1186/2047-783X-16-5-223

10. Been J V., Lugtenberg MJ, Smets E, et al. Preterm Birth and Childhood Wheezing Disorders: A Systematic Review and Meta-Analysis. Lanphear BP, ed. PLoS Med. 2014;11(1):e1001596. doi:10.1371/journal.pmed.1001596

11. Simpson SJ, Turkovic L, Wilson AC, et al. Lung function trajectories throughout childhood in survivors of very preterm birth: a longitudinal cohort study. Lancet Child Adolesc Heal. 2018;2(5):350–359. doi:10.1016/S2352-4642(18)30064-6

12. Treyvaud K, Lee KJ, Doyle LW, Anderson PJ. Very Preterm Birth Influences Parental Mental Health and Family Outcomes Seven Years after Birth. J Pediatr. 2014;164(3):515–521. doi:10.1016/J.JPEDS.2013.11.001

13. Henderson J, Carson C, Redshaw M. Impact of preterm birth on maternal well-being and women’s perceptions of their baby: a population-based survey. BMJ Open. 2016;6(10):e012676. doi:10.1136/bmjopen-2016-012676

14. Carson C, Redshaw M, Gray R, Quigley MA. Risk of psychological distress in parents of preterm children in the first year: evidence from the UK Millennium Cohort Study. BMJ Open. 2015;5(12):e007942. doi:10.1136/bmjopen-2015-007942

15. Chiafery MC, D’Angio CT. Burden of Chronic Lung Disease on the Caregivers. In: Updates on Neonatal Chronic Lung Disease. Elsevier; 2020:317–333. doi:10.1016/B978-0-323-68353-1.00022-1

16. McGrath-Morrow SA, Ryan T, Riekert K, Lefton-Greif MA, Eakin M, Collaco JM. The impact of bronchopulmonary dysplasia on caregiver health related quality of life during the first 2 years of life. Pediatr Pulmonol. 2013;48(6):579–586. doi:10.1002/ppul.22687

17. Singer LT, Fulton S, Kirchner HL, et al. Longitudinal Predictors of Maternal Stress and Coping After Very Low-Birth-Weight Birth. Arch Pediatr Adolesc Med. 2010;164(6):518–524. doi:10.1001/archpediatrics.2010.81

18. Adams M, Bucher H-U. Neonatologie: Ein früher Start ins Leben: Was bringt ein nationales Register? Swiss Med Forum – Schweizerisches Medizin-Forum. 2013;13(03):35–37. doi:10.4414/smf.2013.01397

19. Pedersen ESL, de Jong CCM, Ardura-Garcia C, et al. The Swiss Paediatric Airway Cohort (SPAC). ERJ Open Res. 2018;4(4):00050–02018. doi:10.1183/23120541.00050-2018

20. Medrano GR, Berlin KS, Hobart Davies W. Utility of the PedsQL<>TM<> family impact module: assessing the psychometric properties in a community sample. Qual Life Res. 2013;22(10):2899–2907. doi:10.1007/s11136-013-0422-9

21. Varni JW, Sherman SA, Burwinkle TM, Dickinson PE, Dixon P. The PedsQL<>TM<> Family Impact Module: Preliminary reliability and validity. Health Qual Life Outcomes. 2004;2(1):55. doi:10.1186/1477-7525-2-55

22. Jobe AH, Bancalari E. Bronchopulmonary Dysplasia. Am J Respir Crit Care Med. 2001;163(7):1723–1729. doi:10.1164/ajrccm.163.7.2011060

23. Schlapbach LJ, Adams M, Proietti E, et al. Outcome at two years of age in a Swiss national cohort of extremely preterm infants born between 2000 and 2008. BMC Pediatr. 2012;12(1):1–12. doi:10.1186/1471-2431-12-198/FIGURES/3

24. De Onis M. 4.1 The WHO Child Growth Standards. World Rev Nutr Diet. 2015;113:278–294. doi:10.1159/000360352

25. Verheggen M, Wilson AC, Pillow JJ, Stick SM, Hall GL. Respiratory function and symptoms in young preterm children in the contemporary era. Pediatr Pulmonol. 2016;51(12):1347–1355. doi:10.1002/PPUL.23487

26. Kotecha S, Clemm H, Halvorsen T, Kotecha SJ. Bronchial hyper-responsiveness in preterm-born subjects: A systematic review and meta-analysis. Pediatr Allergy Immunol. 2018;29(7):715–725. doi:10.1111/pai.12957

27. Skromme K, Vollsæter M, Øymar K, Markestad T, Halvorsen T. Respiratory morbidity through the first decade of life in a national cohort of children born extremely preterm. BMC Pediatr. 2018;18(1):102. doi:10.1186/s12887-018-1045-7

28. Pérez-Tarazona S, Rueda Esteban S, García-García ML, et al. Respiratory outcomes of “new” bronchopulmonary dysplasia in adolescents: A multicenter study. Pediatr Pulmonol. 2021;56(5):1205–1214. doi:10.1002/ppul.25226

29. Fawke J, Lum S, Kirkby J, et al. Lung function and respiratory symptoms at 11 years in children born extremely preterm: The EPICure study. Am J Respir Crit Care Med. 2010;182(2):237–245. doi:10.1164/rccm.200912-1806OC

30. Manti S, Galdo F, Parisi GF, et al. Long-term effects of bronchopulmonary dysplasia on lung function: a pilot study in preschool children’s cohort. J Asthma. 2021;58(9):1186–1193. doi:10.1080/02770903.2020.1779289

31. Yammine S, Schmidt A, Sutter O, et al. Functional evidence for continued alveolarisation in former preterms at school age? Eur Respir J. 2016;47(1):147–155. doi:10.1183/13993003.00478-2015

32. Narayanan M, Beardsmore CS, Owers-Bradley J, et al. Catch-up Alveolarization in Ex-Preterm Children. Evidence from 3 He Magnetic Resonance. Am J Respir Crit Care Med. 2013;187(10):1104–1109. doi:10.1164/rccm.201210-1850OC

33. Cazzato S, Ridolfi L, Bernardi F, Faldella G, Bertelli L. Lung function outcome at school age in very low birth weight children. Pediatr Pulmonol. 2013;48(8):830–837. doi:10.1002/PPUL.22676

34. Doyle LW, Faber B, Callanan C, Freezer N, Ford GW, Davis NM. Bronchopulmonary Dysplasia in Very Low Birth Weight Subjects and Lung Function in Late Adolescence. Pediatrics. 2006;118(1):108–113. doi:10.1542/PEDS.2005-2522

35. Hart K, Cousins M, Watkins WJ, Kotecha SJ, Henderson AJ, Kotecha S. Association of early-life factors with prematurity-associated lung disease: prospective cohort study. Eur Respir J. 2022;59(5):2101766. doi:10.1183/13993003.01766-2021

36. Singer LT, Fulton S, Kirchner HL, et al. Parenting Very Low Birth Weight Children at School Age: Maternal Stress and Coping. J Pediatr. 2007;151(5):463–469. doi:10.1016/j.jpeds.2007.04.012

37. Treyvaud K, Lee KJ, Doyle LW, Anderson PJ. Very Preterm Birth Influences Parental Mental Health and Family Outcomes Seven Years after Birth. J Pediatr. 2014;164(3):515–521. doi:10.1016/J.JPEDS.2013.11.001

38. Foronda CL, Kelley CN, Nadeau C, et al. Psychological and Socioeconomic Burdens Faced by Family Caregivers of Children With Asthma: An Integrative Review. J Pediatr Heal Care. 2020;34(4):366–376. doi:10.1016/j.pedhc.2020.02.003

39. Fawcett R, Porritt K, Stern C, Carson-Chahhoud K. Experiences of parents and carers in managing asthma in children. JBI Database Syst Rev Implement Reports. 2019;17(5):793–984. doi:10.11124/JBISRIR-2017-004019

40. Caicedo C. Families With Special Needs Children. J Am Psychiatr Nurses Assoc. 2014;20(6):398–407. doi:10.1177/1078390314561326

41. Hatzmann J, Peek N, Heymans H, Maurice-Stam H, Grootenhuis M. Consequences of caring for a child with a chronic disease. J Child Heal Care. 2014;18(4):346–357. doi:10.1177/1367493513496668

42. Sullivan PW, Ghushchyan V, Navaratnam P, et al. The national burden of poorly controlled asthma, school absence and parental work loss among school-aged children in the United States. J Asthma. 2018;55(6):659–667. doi:10.1080/02770903.2017.1350972

43. Mogensen I, Hallberg J, Ekström S, Bergström A, Melén E, Kull I. Uncontrolled asthma from childhood to young adulthood associates with airflow obstruction. ERJ Open Res. 2021;7(4):00179–02021. doi:10.1183/23120541.00179-2021

44. Tsuneyoshi S, Kawayama T, Sasaki J, et al. Poor Asthma Control in Schoolchildren May Lead to Lower Lung Function Trajectory from Childhood to Early Adulthood: A Japanese Cohort Study. J Asthma Allergy. 2022;Volume 15:885–896. doi:10.2147/JAA.S366453

45. Lodge CJ, Lowe AJ, Allen KJ, et al. Childhood Wheeze Phenotypes Show Less Than Expected Growth in FEV 1 across Adolescence. Am J Respir Crit Care Med. 2014;189(11):1351–1358. doi:10.1164/rccm.201308-1487OC

46. Cano-Garcinuño A, Mora-Gandarillas I, Bercedo-Sanz A, et al. Looking beyond patients: Can parents’ quality of life predict asthma control in children? Pediatr Pulmonol. 2016;51(7):670–677. doi:10.1002/ppul.23336

