## Supplement for "Respiratory morbidity in preschool and school-age children born very preterm and its association with parents’ health-related quality of life and family functioning"

* Shared first authorship

### Supplementary methods

#### Study design and participants

In this cross-sectional study we recruited children born less than 32 weeks' gestation between January 2006 and December 2019, in the greater Zurich area, Switzerland. They were all included in the Swiss Neonatal Network & Follow-Up Group (SwissNeoNet), a nationwide registry containing information about all children born less than 32 weeks' gestation since the year 2000.[1] The postal addresses of all families were retrieved from the neonatology ward of the University Hospital of Zurich and the Children's University Hospital of Zurich. Of 1,697 eligible children, valid postal addresses were obtained for 1,401 of them. Invitation letters were sent out in six rounds between May and December 2021. Parents were invited to complete an online survey for their preterm child as well as for a term born (≥37 weeks' gestation) sibling aged 1 to 18 years, referred as controls hereafter. Families who did not complete the survey within two weeks received a reminder call or a second invitation letter, if the phone number was not available. Parents were given the option to complete a paper version and with translation into English, French and Italian. A voucher for a fun park or a plush toy was offered to the child of each participating family. We finalized data collection in May 2022.

The online survey was completed for 681 children born very preterm and 205 controls. Out of them, we excluded 90 children:

- 10 very preterm children with severe chronic conditions: four with severe cerebral palsy (i.e., need of wheelchair and significant difficulty in managing daily activities), one with trisomy 9, one with trisomy 21, one with spina bifida, one with hydrocephalus, one with arthrogryposis multiplex congenita, and one with long-gap esophageal atresia.
- 5 very preterm children without data for the assessed respiratory symptoms
- 50 very preterm children and 25 controls with a recent COVID-19 infection. A COVID-19 infection was defined based on a positive answer for the two following questions: *Has your child had a coronavirus infection?, Has this infection been confirmed by a test?*

Thus, the study sample consisted of 616 very preterm children (44% of the invited) and 180 controls. The analysis on the association between children's respiratory symptoms and parents' HRQoL was restricted to very preterm children and for families with twins or triplets one child was randomly selected (n=533, Figure S1).

The study was approved by the Ethics Committee of the Canton of Zurich, Switzerland (2020-02396). Filling out the online survey was considered as providing consent, which enabled us to claim an exception from written consent form (in accordance with the Ethics Committee regulation).

#### Covariates

Neonatal characteristics for premature children were retrieved from the SwissNeoNet registry. Gestational age was categorized as 22 to 28 weeks (extremely preterm) and 28 to 31 weeks (very preterm). BPD status was defined as oxygen use for 28 days and its severity was defined based on oxygen dependency at 36 weeks post-menstrual age.[2] Premature children were classified as no-to-mild BPD and moderate-to-severe BPD.[2] Birth weight z-scores and socioeconomic status of the family were defined as previously published.[3]

We collected information on children's sex, age, weight, and height in the online survey. We used height and weight to derive age- and sex-specific body mass index (BMI) z-scores.[4] We also collected information on maternal smoking during pregnancy, parents' nationality (at least one Swiss; both non-Swiss), educational level (high: at least one with preparatory high school to university; low/medium: both up to apprenticeship or professional school), smoking status (at least one current smoker; both non-smokers or ex-smokers) and history of atopy (at least one with history of asthma, eczema or hay fever; both without history of atopy). Finally, we also assessed the number of siblings, the presence of chronic conditions (including minimal to moderate cerebral palsy, heart disease, allergies, diabetes, chronic bowel inflammation, attention deficit disorder, epilepsy, joint disease, depression and anxiety) and who filled in the questionnaire. We classified children's age as 1 to 5 years (preschool age) and 6 to 18 years (school age).

**References**

1. Adams M, Bucher H-U. Neonatologie: Ein früher Start ins Leben: Was bringt ein nationales Register? *Swiss Med Forum – Schweizerisches Medizin-Forum*. 2013;13(03):35-37. doi:10.4414/smf.2013.01397

2. Jobe AH, Bancalari E. Bronchopulmonary Dysplasia. *Am J Respir Crit Care Med*. 2001;163(7):1723-1729. doi:10.1164/ajrccm.163.7.2011060

3. Schlapbach LJ, Adams M, Proietti E, et al. Outcome at two years of age in a Swiss national cohort of extremely preterm infants born between 2000 and 2008. *BMC Pediatr*. 2012;12(1):1-12. doi:10.1186/1471-2431-12-198/FIGURES/3

4. De Onis M. 4.1 The WHO Child Growth Standards. *World Rev Nutr Diet*. 2015;113:278-294. doi:10.1159/000360352

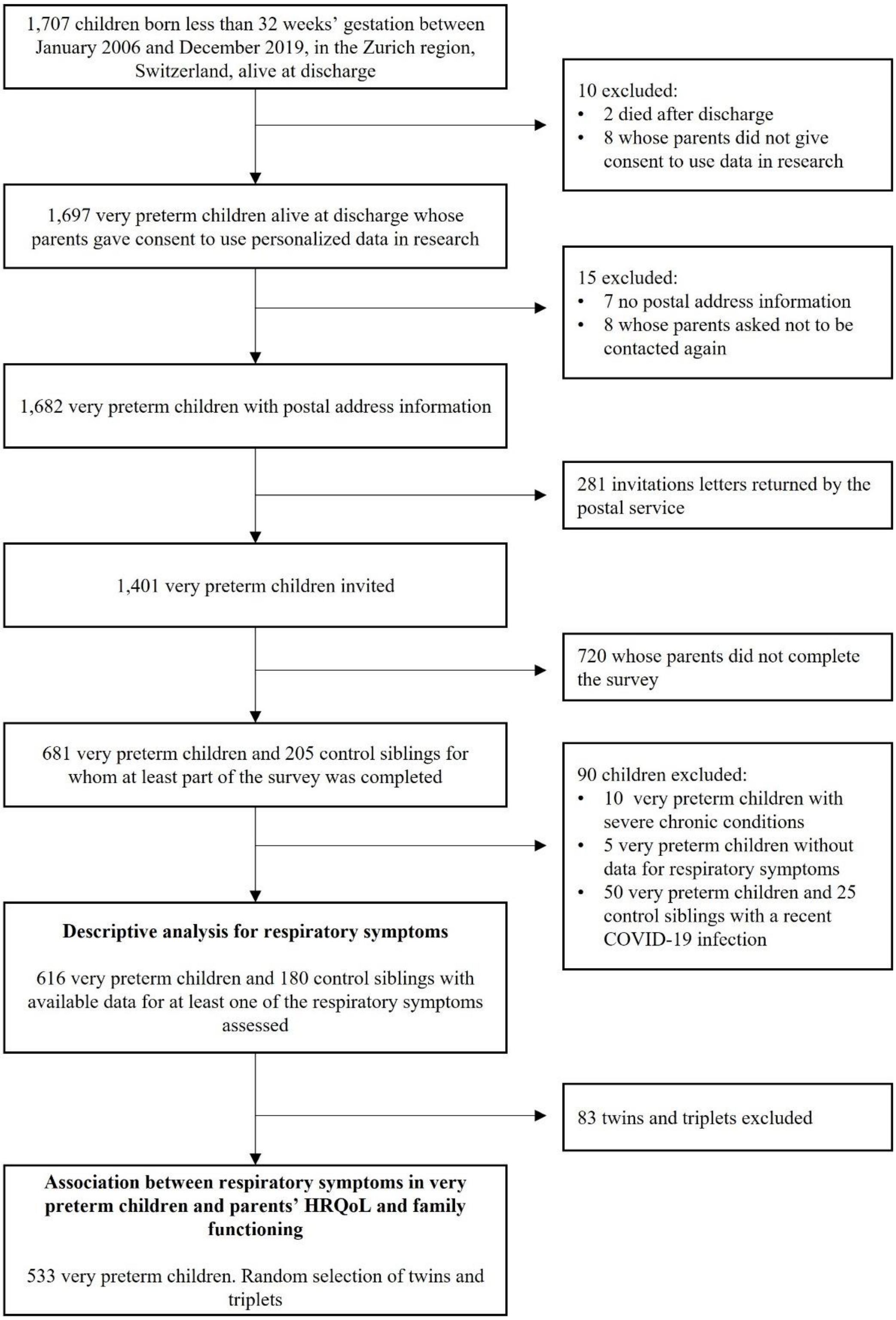

**Figure S1. Flowchart of the study sample**

Abbreviations: HRQoL, health-related quality of life.

**Table S1. Questions used to assess respiratory symptoms**

| Symptoms | Question from the Swiss Paediatric Airway Cohort (SPAC)[1] questionnaire | Positive answer |
|---|---|---|
| **Mild** | *Children were classified as having mild symptoms if they had a positive answer in any of the following four questions* | |
| Breathing difficulties during exertion | Does your child sometimes have difficulty breathing during physical exertion? | Yes |
| Cough without cold | Does your child cough even if he or she does not have a cold? | "Yes, often" or "yes, sometimes" |
| Nocturnal cough past 12 months | In the last 12 months, has your child ever had a dry, irritating cough at night, even though he or she did not have a cold or bronchitis? | Yes |
| Wheezing past 12 months | Has your child had whistling or wheezing breathing in the last 12 months? | Yes |
| **Moderate-severe** | *Children were classified as having moderate-severe symptoms if they had a positive answer in any of the following two questions* | |
| Emergency visits or hospitalization past 12 months | In the last 12 months, has your child ever been treated for coughing, wheezing or asthma?<br>• In an emergency at the pediatrician or family doctor<br>• In the emergency ward of a hospital (without overnight stay)<br>• In-patient at a hospital (i.e., your child has stayed overnight one or more times) | Yes |
| Inhalation therapy past 12 months | Has your child inhaled a spray or powder preparation for respiratory problems in the last 12 months? | Yes |

[1]Reference: Pedersen ESL, de Jong CCM, Ardura-Garcia C, et al. The Swiss Paediatric Airway Cohort (SPAC). *ERJ Open Res*. 2018;4(4):00050-02018.

**Table S2. Differences in children's and parents' characteristics between observed and imputed datasets used for multivariate analyses (n=533)**

| | % Missing | Observed data | Imputed data |
|---|---|---|---|
| **Children's characteristics** | | | |
| Female sex | 0% | 45% | 45% |
| Gestational age categories | 0% | | |
| 28 to 31 weeks | | 66% | 66% |
| 22 to 28 weeks | | 34% | 34% |
| Age categories | 0% | | |
| 1 to 5 years | | 40% | 40% |
| 6 to 16 years | | 60% | 60% |
| Chronic conditions | 4.5% | 20% | 20% |
| Number of siblings | 3.8% | | |
| None | | 27% | 27% |
| One | | 50% | 50% |
| Two or more | | 22% | 22% |
| **Parents' characteristics** | | | |
| Smoking, at least one current smoker | 5.1% | 22% | 22% |
| Nationality, at least one Swiss | 3.4% | 81% | 81% |
| Education, at least one high level | 5.4% | 73% | 73% |
| History of atopy, at least one | 5.3% | 59% | 59% |
| Who completed the questionnaire | 0.8% | | |
| Mother alone | | 68% | 68% |
| Mother and father | | 21% | 21% |
| Father alone | | 11% | 11% |
| **Combined variable for respiratory symptoms** | 20.3% | | |
| None | | 54% | 56% |
| Mild | | 25% | 24% |
| Moderate-severe | | 21% | 19% |
| **PedsQL FIM scores** | | | |
| Total score | 5.1% | 98.6 (88.9; 100.0) | 98.6 (88.9; 100.0) |
| Parent HRQoL summary score | 4.5% | 98.8 (88.8; 100.0) | 98.8 (88.8; 100.0) |
| Family functioning summary score | 6.8% | 100.0 (84.4; 100.0) | 100.0 (84.4; 100.0) |

Data are presented as median ($P_{25}$; $P_{75}$) or %. Some percenatges do not add to 100% because of rounding.

**Table S3. 1Characteristics of participants and non-participants**

| | Participants | | |
|---|---|---|---|
| **Characteristic** | **No, n = 1,081** | **Yes, n = 616** | **p-value[1]** |
| Female sex | 493/1,081 (46%) | 275/616 (45%) | 0.702 |
| Age (years) | 7.9 (4.7; 11.7) | 7.0 (4.0; 10.9) | 0.003 |
| Gestational age (weeks) | 30 (28; 31) | 29 (27; 30) | <0.001 |
| Gestational age, categories | | | <0.001 |
| 28-32 weeks | 819/1,081 (76%) | 404/616 (66%) | |
| <28 weeks | 262/1,081 (24%) | 212/616 (34%) | |
| Birth weight (g) | 1,260 (980; 1,520) | 1,140 (858; 1,450) | <0.001 |
| Birth weight, z-score | 0.0 (-0.5; 0.5) | -0.1 (-0.6; 0.4) | 0.060 |
| Moderate-to-severe BPD | 104/1,081 (10%) | 99/615 (16%) | <0.001 |
| Socioeconomic status score | 6 (4; 8) | 5 (3; 6) | <0.001 |

Data are presented as median ($P_{25}$; $P_{75}$) or n/N (%).[1]Pearson's Chi-squared test; Wilcoxon rank sum test

Abbreviations: BPD, bronchopulmonary dysplasia.

**Table S4. Between group comparisons for mild and moderate-severe respiratory symptoms in preschool-age children**

| | VP no BPD *vs.* C | | VP BPD *vs.* C | | VP BPD *vs.* VP no BPD | |
|---|---|---|---|---|---|---|
| | **OR [95% CI]** | **p-value** | **OR [95% CI]** | **p-value** | **OR [95% CI]** | **p-value** |
| **Mild symptoms** | 3.1 [1.6 to 6.1] | <0.001 | 4.1 [1.8 to 9.6] | <0.001 | 1.3 [0.7 to 2.6] | 0.365 |
| Breathing difficulties during exertion | 6.3 [1.2 to 114.9] | 0.077 | 10.5 [1.8 to 200] | 0.031 | 1.7 [0.6 to 4.2] | 0.292 |
| Cough without cold | 2.0 [0.8 to 5.4] | 0.150 | 3.7 [1.4 to 11.4] | 0.013 | 1.9 [0.9 to 3.9] | 0.082 |
| Nocturnal cough past 12 months | 5.5 [1.9 to 23.4] | 0.006 | 6.6 [1.9 to 30.6] | 0.006 | 1.2 [0.5 to 2.5] | 0.642 |
| Wheezing past 12 months | 1.9 [0.9 to 5.0] | 0.136 | 2.3 [0.8 to 6.7] | 0.117 | 1.2 [0.5 to 2.5] | 0.673 |
| **Moderate-severe symptoms** | 1.9 [0.9 to 4.2] | 0.103 | 2.4 [1.0 to 6.1] | 0.053 | 1.3 [0.6 to 2.5] | 0.472 |
| Emergency visits or hospitalization past 12 months | 2.2 [0.9 to 6.0] | 0.104 | 3.2 [1.2 to 9.8] | 0.030 | 1.5 [0.7 to 3.1] | 0.301 |
| Inhalation therapy past 12 months | 1.4 [0.6 to 3.5] | 0.407 | 2.2 [0.8 to 6.0] | 0.124 | 1.5 [0.7 to 3.2] | 0.282 |

Between group comparisons are presented as unadjusted OR (95% CI).

Abbreviations: BPD, bronchopulmonary dysplasia; C, controls; CI, confidence interval; OR, odds ratio; VP no BPD, very preterm with no-to-mild BPD; VP BPD, very preterm with moderate-to-severe BPD.

**Table S5. Between group comparisons for mild and moderate-severe respiratory symptoms in school-age children**

| | **VP no BPD *vs.* C** | | **VP BPD *vs.* C** | | **VP BPD *vs.* VP no BPD** | |
|---|---|---|---|---|---|---|
| | **OR [95% CI]** | **p-value** | **OR [95% CI]** | **p-value** | **OR [95% CI]** | **p-value** |
| **Mild symptoms** | 1.4 [0.9 to 2.3] | 0.188 | 2.0 [0.9 to 4.2] | 0.090 | 1.4 [0.7 to 2.8] | 0.333 |
| Breathing difficulties during exertion | 2.7 [1.2 to 7.4] | 0.026 | 3.4 [1.1 to 11.3] | 0.037 | 1.2 [0.5 to 2.9] | 0.618 |
| Cough without cold | 0.9 [0.5 to 1.6] | 0.736 | 0.8 [0.3 to 2.0] | 0.678 | 0.9 [0.4 to 2.0] | 0.814 |
| Nocturnal cough past 12 months | 1.2 [0.6 to 2.6] | 0.567 | 1.3 [0.4 to 4.0] | 0.602 | 1.1 [0.4 to 2.8] | 0.859 |
| Wheezing past 12 months | 2.0 [0.9 to 5.4] | 0.138 | 1.3 [0.3 to 5.1] | 0.730 | 0.7 [0.2 to 1.9] | 0.494 |
| **Moderate-severe symptoms** | 1.5 [0.7 to 3.3] | 0.275 | 2.4 [0.8 to 6.5] | 0.093 | 1.6 [0.6 to 3.5] | 0.285 |
| Emergency visits or hospitalization past 12 months | 1.5 [0.6 to 5.5] | 0.443 | 2.7 [0.6 to 12.1] | 0.169 | 1.8 [0.5 to 5.1] | 0.326 |
| Inhalation therapy past 12 months | 1.9 [0.9 to 4.8] | 0.137 | 3.7 [1.2 to 11.2] | 0.019 | 1.9 [0.8 to 4.4] | 0.126 |

Between group comparisons are presented as unadjusted OR (95% CI).

Abbreviations: BPD, bronchopulmonary dysplasia; C, controls; CI, confidence interval; OR, odds ratio; VP no BPD, very preterm with no-to-mild BPD; VP BPD, very preterm with moderate-to-severe BPD.

**Table S6. Combined variable for respiratory symptoms, stratified by age and BPD categories**

| **Preschool-age children** | **C, n = 67** | **VP no BPD, n = 191** | **VP BPD, n = 55** | **p-value[1]** |
|---|---|---|---|---|
| **Respiratory symptoms** | | | | 0.007 |
| None | 44/62 (71%) | 73/154 (47%) | 17/45 (38%) | |
| Mild | 8/62 (13%) | 38/154 (25%) | 12/45 (27%) | |
| Moderate-severe | 10/62 (16%) | 43/154 (28%) | 16/45 (36%) | |
| **School-age children** | **C, n = 113** | **VP no BPD, n = 325** | **VP BPD, n = 44** | |
| **Respiratory symptoms** | | | | 0.284 |
| None | 71/103 (69%) | 163/268 (61%) | 19/34 (56%) | |
| Mild | 22/103 (21%) | 65/268 (24%) | 7/34 (21%) | |
| Moderate-severe | 10/103 (10%) | 40/268 (15%) | 8/34 (24%) | |

Data are presented as n/N (%).[1]Group comparisons were tested using Fisher's exact test.

Abbreviations: BPD, bronchopulmonary dysplasia; C, controls; VP no BPD, very preterm with no-to-mild BPD; VP BPD, very preterm with moderate-to-severe BPD.

**Table S7. PedsQL Family Impact Module Scores according to age categories**

| Summary scores | Overall, n = 533 | Preschool-age children, n = 212 | School-age children, n = 321 | p-value[1] |
|---|---|---|---|---|
| Total score | 98.6 (88.9; 100.0) | 97.6 (82.6; 100.0) | 98.9 (91.7; 100.0) | 0.045 |
| Parent HRQoL Summary Score | 98.8 (88.8; 100.0) | 97.5 (85.0; 100.0) | 100.0 (91.2; 100.0) | 0.026 |
| Family functioning Summary Score | 100.0 (84.4; 100.0) | 100.0 (81.2; 100.0) | 100.0 (87.5; 100.0) | 0.216 |

Data are presented as median ($P_{25}$; $P_{75}$). [1]Wilcoxon rank sum test

Abbreviations: HRQoL, health-related quality of life.

**Table S8. Adjusted associations between respiratory symptoms in very preterm children and parents' HRQoL and family functioning**

| | Total Score | | Parent HRQoL | | Family Functioning | |
|---|---|---|---|---|---|---|
| | **Coef. [95% CI]** | **p-value** | **Coef. [95% CI]** | **p-value** | **Coef. [95% CI]** | **p-value** |
| **Overall sample** | | | | | | |
| None | Ref. | | Ref. | | Ref. | |
| Mild | -3.9 [-6.6 to -1.1] | 0.006 | -4.4 [-7.2 to -1.7] | 0.002 | -4.8 [-8.4 to -1.1] | 0.010 |
| Moderate-severe | -8.2 [-11.2 to -5.2] | <0.001 | -7.9 [-10.9 to -4.9] | <0.001 | -9.1 [-13.1 to -5.1] | <0.001 |
| **Preschool-age children** | | | | | | |
| None | Ref. | | Ref. | | Ref. | |
| Mild | -3.3 [-7.9 to 1.3] | 0.153 | -4.2 [-8.8 to 0.4] | 0.075 | -3.3 [-9.0 to 2.4] | 0.252 |
| Moderate-severe | -7.2 [-11.5 to -2.9] | 0.001 | -7.6 [-12.0 to -3.1] | 0.001 | -8.2 [-13.5 to -2.8] | 0.003 |
| **School-age children** | | | | | | |
| None | Ref. | | Ref. | | Ref. | |
| Mild | -5.0 [-8.5 to -1.5] | 0.006 | -5.4 [-8.9 to -1.8] | 0.003 | -6.5 [-11.4 to -1.6] | 0.010 |
| Moderate-severe | -9.2 [-13.4 to -5.0] | <0.001 | -8.2 [-12.4 to -4.0] | <0.001 | -10.1 [-15.9 to -4.2] | 0.001 |

Models were adjusted for child's sex, age category, gestational age category, presence of chronic diseases, number of siblings, parents' nationality, educational level, smoking status, family history of atopy and for who filled in the survey. Models stratified by age categories were adjusted for the same variables, except age.

Abbreviations: CI, confidence interval; Coef., regression coefficient; HRQoL, health-related quality of life; Ref., reference.

**Table S9. Adjusted associations between respiratory symptoms in very preterm children and parents' HRQoL and family functioning: Excluding extreme values (i.e., below the first percentile)**

| | Total Score | | Parent HRQoL | | Family Functioning | |
|---|---|---|---|---|---|---|
| | **Coef. [95% CI]** | **p-value** | **Coef. [95% CI]** | **p-value** | **Coef. [95% CI]** | **p-value** |
| **Overall sample** | | | | | | |
| None | Ref. | | Ref. | | Ref. | |
| Mild | -3.9 [-6.5 to -1.3] | 0.004 | -4.3 [-6.8 to -1.7] | 0.001 | -4.8 [-8.3 to -1.4] | 0.006 |
| Moderate-severe | -8.3 [-11.1 to -5.5] | <0.001 | -7.8 [-10.6 to -5] | <0.001 | -9.4 [-13.2 to -5.6] | <0.001 |
| **Preschool-age children** | | | | | | |
| None | Ref. | | Ref. | | Ref. | |
| Mild | -3.7 [-7.9 to 0.5] | 0.086 | -4.5 [-8.8 to -0.3] | 0.038 | -3.7 [-9.4 to 2] | 0.203 |
| Moderate-severe | -7.0 [-11.1 to -3.0] | 0.001 | -7.2 [-11.3 to -3.1] | 0.001 | -8.4 [-13.8 to -3] | 0.003 |
| **School-age children** | | | | | | |
| None | Ref. | | Ref. | | Ref. | |
| Mild | -4.7 [-8 to -1.3] | 0.007 | -4.8 [-8.1 to -1.5] | 0.005 | -6.1 [-10.6 to -1.6] | 0.008 |
| Moderate-severe | -9.8 [-13.8 to -5.8] | <0.001 | -8.7 [-12.6 to -4.7] | <0.001 | -10.5 [-15.9 to -5.2] | <0.001 |

Models were adjusted for child's sex, age category, gestational age category, presence of chronic diseases, number of siblings, parents' nationality, educational level, smoking status, family history of atopy and for who filled in the survey. Models stratified by age categories were adjusted for the same variables, except age.

Abbreviations: CI, confidence interval; Coef., regression coefficient; HRQoL, health-related quality of life; Ref., reference.

**Table S10. Adjusted associations between respiratory symptoms in very preterm children and parents' HRQoL and family functioning: Using the observed data**

| | Total Score | | Parent HRQoL | | Family Functioning | |
|---|---|---|---|---|---|---|
| | **Coef. [95% CI]** | **p-value** | **Coef. [95% CI]** | **p-value** | **Coef. [95% CI]** | **p-value** |
| **Overall sample** | | | | | | |
| None | Ref. | | Ref. | | Ref. | |
| Mild | -3.5 [-6.5 to -0.5] | 0.021 | -3.9 [-6.9 to -0.8] | 0.014 | -4.1 [-8.1 to -0.1] | 0.043 |
| Moderate-severe | -8.4 [-11.7 to -5.1] | <0.001 | -7.9 [-11.3 to -4.5] | <0.0001 | -9.3 [-13.8 to -4.9] | <0.0001 |
| **Preschool-age children** | | | | | | |
| None | Ref. | | Ref. | | Ref. | |
| Mild | -2.6 [-7.8 to 2.5] | 0.312 | -3.0 [-8.4 to 2.4] | 0.271 | -1.7 [-8.1 to 4.6] | 0.59 |
| Moderate-severe | -8.3 [-13.3 to -3.3] | 0.001 | -8.2 [-13.5 to -3.0] | 0.002 | -9.5 [-15.7 to -3.2] | 0.003 |
| **School-age children** | | | | | | |
| None | Ref. | | Ref. | | Ref. | |
| Mild | -5.5 [-9.3 to -1.7] | 0.005 | -5.9 [-9.7 to -2.1] | 0.003 | -6.8 [-12.1 to -1.4] | 0.014 |
| Moderate-severe | -8.3 [-12.7 to -3.9] | <0.001 | -7.2 [-11.6 to -2.7] | 0.002 | -9.0 [-15.2 to -2.7] | 0.005 |

Models were adjusted for child's sex, age category, gestational age category, presence of chronic diseases, number of siblings, parents' nationality, educational level, smoking status, family history of atopy and for who filled in the survey. Models stratified by age categories were adjusted for the same variables, except age.

Abbreviations: CI, confidence interval; Coef., regression coefficient; HRQoL, health-related quality of life; Ref., reference.

**Table S11. Adjusted associations between respiratory symptoms in very preterm children and parents' HRQoL and family functioning: Using 100 different random samples for the selection of twins and triplets**

| | Total Score | Parent HRQoL | Family Functioning |
|---|---|---|---|
| | **Coef. [95% CI]** | **Coef. [95% CI]** | **Coef. [95% CI]** |
| **Overall sample** | | | |
| None | Ref. | Ref. | Ref. |
| Mild | -3.3 [-4.2 to -2.6] | -3.7 [-4.5 to -3.0] | -4.1 [-5.1 to -3.0] |
| Moderate-severe | -8.2 [-9.5 to -6.9] | -7.8 [-8.6 to -6.9] | -9 [-10.6 to -7.5] |
| **Preschool-age children** | | | |
| None | Ref. | Ref. | Ref. |
| Mild | -3.1 [-4.3 to -1.9] | -3.5 [-4.5 to -2.4] | -2.2 [-4.0 to -0.3] |
| Moderate-severe | -9.1 [-11.1 to -7.1] | -8.9 [-10.4 to -7.2] | -10.3 [-12.7 to -8.3] |
| **School-age children** | | | |
| None | Ref. | Ref. | Ref. |
| Mild | -4.6 [-5.5 to -3.6] | -4.9 [-5.9 to -3.8] | -5.9 [-7.2 to -4.6] |
| Moderate-severe | -7.3 [-8.9 to -5.5] | -6.4 [-8.0 to -4.8] | -8.0 [-10.0 to -5.9] |

Presented values correspond to the average of the regression coefficients in the 100 random samples. 95% CI for these estimates were calculated using the quantile function.

Models were adjusted for child's sex, age category, gestational age category, presence of chronic diseases, number of siblings, parents' nationality, educational level, smoking status, family history of atopy and for who filled in the survey. Models stratified by age categories were adjusted for the same variables, except age.

Abbreviations: CI, confidence interval; Coef., regression coefficient; HRQoL, health-related quality of life; Ref., reference.
